# The *16p11.2* microdeletion enhances gene expression variability between human IPSC derived forebrain interneuron progenitor cells in culture

**DOI:** 10.64898/2026.05.21.26353723

**Authors:** Yifei Yang, Idoia Quintana Urzainqui, Thomas Pratt

**Affiliations:** Simons Initiative for the Developing Brain; Edinburgh Medical School Biomedical Sciences, Institute for Neuroscience and Cardiovascular Research, University of Edinburgh, United Kingdom; Developmental Biology Unit, European Molecular Biology Laboratory (EMBL), Heidelberg, Germany

**Author notes:** First author equal contribution (Yifei Yang, Idoia Quintana Urzainqui). Corresponding authour.

**Keywords:** Development, neurodevelopmental conditions, *16p11.2* syndrome, ventral telencephalon, single cell RNA sequencing, regulon, gene regulatory network, heterogeneity, induced pluripotent stem cells

## Abstract

The 574 kilobase pair *16p11.2* microdeletion raises a person’s odds for neurodevelopmental and energy balance conditions, particularly autism and obesity. There is considerable clinical heterogeneity and how much this reflects genetic versus environmental or stochastic factors is unclear. Forebrain interneurons originate from progenitors residing in the ventricular zone of the foetal ventral telencephalon and their perturbation is implicated in a number of *16p11.2* phenotypes prompting investigation of how the *16p11.2* microdeletion impacts their development. We differentiate human induced pluripotent stem cells (IPSCs), isogenic except for heterozygous *16p11.2* microdeletion to minimise confounding effects of genetic background, to ventral telencephalic interneuron progenitor fate in 2D culture and use single cell RNA sequencing to obtain single cell transcriptome populations for comparative bioinformatics. Hundreds of transcripts are differentially expressed and many associate with cell signalling, chromatin, neurodevelopmental conditions including autism, and obesity. Pertinently, we find that transcript level variation is significantly greater in *16p11.2* heterozygous progenitors than their isogenic wild type counterparts and this holds for sets of genes comprising regulons, gene-sets functionally connected by transcription factor regulation, and for randomly selected gene-sets indicating that the *16p11.2* locus itself has a genome-wide property in stabilising transcription between cells. Regulons with greatest increased variation in *16p11.2* heterozygous progenitors exhibit strong enrichment for cell cycle related genes, resonating with our earlier finding of increased cell cycle variability between *16p11.2* heterozygous organoids, and many are regulated by transcription factors associated with autism and/or obesity enforcing the idea that unusual transcriptional variation itself contributes to phenotypes.

## INTRODUCTION

Copy number variation (CNV) of the 574 kilobase pair (kbp) *16p11.2* locus in the region BP4-BP5 on the short arm of human chromosome 16 is the genetic risk factor for *16p11.2* syndrome. In humans the *16p11.2* locus is flanked by 150kbp repeat sequences and misaligned meiotic homologous recombination generates reciprocal microdeleted and microduplicated *16p11.2* alleles allowing for *16p11.2* CNVs to arise *de novo* at relatively high frequency (D’Angelo et al., 2016; Nuttle et al., 2016). The *16p11.2* locus is a dosage sensor with deviation from a dosage of two *16p11.2* alleles, heterozygosity for the *16p11.2* microdeletion or microduplication, each associating with *16p11.2* neuropsychiatric, anatomical, and energy balance phenotypes (Jacquemont et al., 2011; Maillard et al., 2016; Weiss et al., 2008). Harbouring the *16p11.2* microdeletion increases the odds (odds ratio (OR)) of developing obesity (BMI>30 kg/m^2^) 43-fold, autism 40-fold, any psychiatric disorder 9-fold and ADHD 4-fold (Niarchou et al., 2019; Walters et al., 2010). Most *16p11.2* microdeletion carriers present with more than one symptom with overall ∼70% being obese and/or 15-25% exhibiting autism, epilepsy, intellectual disability, and/or macrocephally (Chung et al., 2021; Rein and Yan, 2020). The variable penetrance and expressivity of the *16p11.2* microdeletion between different carrier individuals has hitherto been attributed to genetic variation outwith the *16p11.2* locus and environmental factors including the maternal environment pre-natally (Girirajan et al., 2012; Hudac et al., 2020). In non-*16p.11.2* contexts the odds of developing obesity associated with monogenic risk factors is modified by polygenic risk (Akbari et al., 2021). Overall it remains an open question if the *16p11.2* locus itself contributes to clinical variation.

Studies using human and animal models have identified *16p11.2* neuronal phenotypes at various stages of development including differential gene expression, cell proliferation, signalling, cell and tissue anatomy, electrophysiology, and behaviour (Antoine et al., 2019; Arbogast et al., 2016; Blumenthal et al., 2014; Connacher et al., 2022; Deshpande et al., 2017; Fetit et al., 2023; Horev et al., 2011; Migliavacca et al., 2015; Pucilowska et al., 2018, 2015; Qiu et al., 2019; Qureshi et al., 2014; Roth et al., 2020; Sundberg et al., 2021; Urresti et al., 2021; Yang et al., 2023). Reduced dosage of individual *16p11.2* genes including *ALDOA*, *KCTD13*, *MAPK3*, *TAOK2, and MVP* have each been reported to impact the proliferation and/or differentiation of neural progenitors (Blaker-Lee et al., 2012; de Anda et al., 2012; Escamilla et al., 2017; Golzio et al., 2012; Kizner et al., 2020; Pucilowska et al., 2012; Richter et al., 2019; Yadav et al., 2017). How the *16p11.2* locus acts, and the extent to which simultaneously reduced dosage of *16p11.2* genes synergise to produce microdeletion phenotypes, is not fully understood.

GABAergic inhibitory neurons (INs or interneurons) form functional circuits with excitatory neurons in the cerebral cortex and subcortically and shifting the balance between excitation and inhibition has long been hypothesised to contribute to autism and its co-occurring conditions (Antoine et al., 2019; Bozzi et al., 2018; Nelson and Valakh, 2015; Puts et al., 2017; Rapanelli et al., 2017; Robertson et al., 2016; Rubenstein and Merzenich, 2003). IN progenitors reside in the ventricular zone of the ganglionic eminences in the ventral telencephalon and subsequently differentiate and migrate to their target areas. In developing human embryos many genes in the *16p11.2* locus are expressed in IN progenitors suggesting they may be vulnerable to altered *16p11.2* dosage (Morson et al., 2021; Shi et al., 2021; Yu et al., 2021). Consistent with this hypothesis we recently showed that isogenic human IPSC derived IN progenitors in *16p11.2^+/-^* organoids exhibit an extended cell-cycle and accelerated maturation and, intriguingly, increased inter-organoid variability in these and other phenotypes despite being genetically homogenous suggesting that the *16p11.2* locus itself regulates variability in this context (Fetit et al., 2023)

Here we use single cell RNA sequencing (scRNAseq) to compare the transcriptomes of IN progenitor populations generated *in vitro* in 2D culture from human IPSCs which are isogenic except for whether or not they are heterozygous for the *16p11.2* microdeletion (Fetit et al., 2020; Yan Liu et al., 2013; Tai et al., 2016). In line with other studies on other *16p11.2^+/-^* cell types we find hundreds of genes are dysregulated at relatively modest levels in *16p11.2^+/-^* IN progenitors. We observe a general increased cell-cell variability in gene expression between isogenic *16p11.2^+/-^* IN progenitor cells suggesting that the *16p11.2* locus normally has a global stabilising effect on gene expression, including on gene sets linked to the cell cycle and *16p11.2* symptoms.

## METHODS

### DNA SNP microarray analysis

We used Illumina HumanCytoSNP-12 v2.1 array analysis to assess genomic structure in the 3 deletion lines (DELD5, DELH7, and DELC5) and the 2 control lines (FACS52 and FACS53) used for this study and their ancestral GM8 line.

### *In vitro* differentiation of hIPSC lines into INs

The hIPSC lines used for this study were CRISPR/Cas9 engineered from the parental GM8 hIPSC line to recapitulate the genomic architecture of the *16p11.2* microdeletion in patients (*16p11.2^+/-^*) or isogenic controls derived from the same GM8 parental line without CRISPR/Cas9 engineering (*16p11.2^+/+^*) (Tai et al., 2016). For single cell RNA sequencing three *16p11.2^+/-^* hIPSC lines each produced by a different targeting event (DELD5, DELC5, and DELH7) were each used to generate a separate culture while one isogenic *16p11.2^+/+^* control line (FACS5.2) was used to generate one culture and another (FACS5.3) was used to generate two cultures. We refer to the *CON* (*16p11.2^+/+^*) cultures as C1-3 and the *DEL* (*16p11.2^+/-^*) cultures as D1-3: C1=FACS5.2; C2=FACS5.3; C3=FACS5.3; D1=DELD5; D2=DELC5, and D3=DELH7. Differentiation of IPSCs was performed as described by (Y Liu et al., 2013) (S2A). For each sample hiPSCs were grown in maintenance medium composed by a 1:1 mix of mTeSR^TM^1 (Stem Cell Technologies) and Essential 8 TM Medium (Thermo Fisher Scientific). hiPSCs were differentiated into INs based on the 2D protocol described in day 0, hiPSCs were lifted and plated in non-adherent plates to allow the formation of embryoid bodies (EBs). On day 4, pluripotency media was replaced by Neural Induction Media (NIM, (Liu et al., 2013)). On day 7, EBs were plated in matrigel-coated plates (Matrigel Matrix, Corning) and allowed to form rosette structures. On day 10, the Shh agonist purmorphamine (Stem Gent, 1.5µM) was added to the media to induce GABAergic differentiation. On day 16, rosettes were manually selected, lifted with a 10ul pipette tip and transferred to a non-adherent plates to allow the formation of neurospheres. B27 supplement (1:50, without Vitamin A, Gibco) was added to the media. On day 26, neurospheres were incubated in accutase solution for 5 minutes at 37°C and dissociated. Single neurons were plated in 24 well plates coated with PLO (Poly-L-ornithine, Sigma-Aldrich) and laminin at a density of 250000 cells per well in Neuronal Differentiation medium containing 1mM cAMP and 100µg/ml of BDNF, GDNF and IGF1. Neurons were fed every three days and left to mature until day 80.

### Immunofluorescence

Cultures were fixed in 4% paraformaldehyde (PFA) for 30 min to 2 h followed by permeabilization in 0.1% Triton X-100 diluted in PBS (PBST) and blocked in 10% donkey or goat serum in PBST for 30 min at room temperature. Then incubated overnight at 4°C with primary antibodies and PBST was used to wash off the primary antibodies followed by incubation with fluorescent conjugated secondary antibodies in blocking solution for 1 h. Finally, nuclei were visualised with DAPI (Hoechst 33342, BD Pharmingen 561908, 5µg/ml) and mounted for microscopy under coverslips using Vectashield Hardset (Vector Labs) for imaging on an epifluorescence microscope. Primary antibodies: FOXG1 (Rabbit, Abcam ab18259, diluted 1/100) and TUJ1 (Mouse, Biolegend 801203 diluted 5µl/ million cells) in blocking solution. Secondary antibodies: Anti-Rabbit-488 (Goat, Alexa Fluor 488, Abcam ab150077 dilution 1/200) and Anti-mouse-647 (Goat, Alexa Fluor 647, Abcam ab150115, 1/200)

### cDNA library preparation and single cell RNA sequencing

Cultures were dissociated to single cell suspension using Accutase. Cells were checked for viability by Trypan Blue staining and cell density estimated in an automated cell counter (Countess, ThermoFisher). Multiplex i7 Barcoded cDNA libraries were prepared for single cell RNA sequencing using 10X Chromium Next GEM v3.1 Single Cell 3’ Library and Gel Bead Kit and loaded onto the 10x Chromium instrument through a Next GEM v3.1 Chip G. We loaded 6 samples in two different runs. In the first run 3 samples from three separate lines were loaded (C1, D1, and D2) in independent channels at a density of 10^6^ cells/ml. In the second run, 3 samples from two separate lines were loaded (C2, C3, and D3) in independent channels. Libraries were constructed following the manufacturer’s protocol. Bioanalyzer traces were assessed for RNA quality and all samples run on one lane on a NovaSeq S1 at 750M read pairs. Multiplex i7 Barcoded cDNA libraries were prepared for single cell RNA sequencing using 10X Chromium Next GEM v3.1 Single Cell 3’ Library and Gel Bead Kit and loaded onto a Next GEM v3.1 Chip G for 10x sequencing.

### Single-Cell RNA-Seq Data Processing and Clustering

CellRanger (ver. 6.0) was used to map fastq files to the human genome assembly version GRCh38. Raw count matrix was processed and analyzed using the *Seurat* R package (ver. 4.3.0). Quality control was performed to filter low-quality cells. Cells were removed if they met any of the following criteria: (1) gene numbers less than 800, (2) gene numbers greater than 10,000, (3) UMI greater than 200,000, (4) percentage of mitochondrial RNA UMIs greater than 10%, (5) percentage of ribosome RNA UMIs greater than 40%, and (6) percentage of red cell genes (“HBA1”,“HBA2”,“HBB”,’HBD’,’HBE1’,’HBG1’,’HBG2’,’HBM’,’HBQ1’,’HBZ’) greater than 0.1%. Following quality control, data were normalized using the *NormalizeData* function, and top 2000 highly variable features (HVGs) were identified using the *FindVariableFeatures* method. Dimensionality reduction was performed via Principal Component Analysis (PCA), followed by non-linear dimensionality reduction using Uniform Manifold Approximation and Projection (UMAP) for visualization. Graph-based clustering was performed using the *FindNeighbors* and *FindClusters* functions with the resolution 0.8. Elimination of batch effects between samples was achieved by Harmony (ver. 0.1.0).

### Cell Type Annotation

Cell types were annotated based on the expression of canonical marker genes. To validate these annotations, a heatmap of the top marker genes for each cluster was generated using the *ggplot* R package (ver. 3.4.4). Clusters were assigned to specific cell types by comparing marker expression profiles against known cell-type signatures.

### Differential Expression Analysis via Pseudo-bulk Aggregation

To identify robust differentially expressed genes (DEGs) between conditions, we performed pseudo-bulk analysis. Single-cell gene expression counts were aggregated to the sample level by summing the counts for each gene across all cells within the same biological replicate and cell type. This generated a “pseudo-bulk” count matrix, which was then used as input for *DESeq2* (ver. 1.34.0). Differential expression testing was performed using a standard likelihood ratio test within the DESeq2 framework. Genes with an adjusted *P*-value < 0.05 were considered as statistically significant DEGs.

### Biological function of DEGs

To investigate the biological functions of the DEGs, gene ontology (GO) annotation and pathway enrichment analysis of the DEGs were carried out by using **Metascape** web tool (http://metascape.org) (Zhou et al., 2019) with the ontology sources of GO Biological Processes, KEGG Pathway, Reactome, Gene Sets, and WikiPathways Pathway. All genes in the human genome were used as the background. Disease enrichment analysis was performed using the **DisGeNET** platform. The list of differentially expressed genes was tested for enrichment against known disease-associated gene sets. Statistical significance was assessed using a hypergeometric test, with Benjamini-Hochberg correction for multiple testing (adjusted P-value < 0.05). Gene-set enrichment analysis (GSEA) was performed to identify enrichment of terms associated with biological processes (Subramanian et al., 2005).

### Genetic risk (BETA) for Autism and Obesity (BMI)tprat

To evaluate the disease risk associated with dysregulation of specific genes we integrated differential expression data with genetic association metrics for Body Mass Index (BMI) and Autism Spectrum Disorder (ASD). BMI-associated risk single nucleotide polymorphisms (SNPs) were acquired from UK Biobank genome-wide association study (GWAS) summary statistics (Sudlow et al., 2015) and variants were mapped to genes within a 33.5 kb flanking window to extract corresponding effect sizes BETA (β) (Reynolds, R., 2022; Available at: https://doi.org/10.5281/zenodo.6127446) which were then averaged for each gene. ASD risk genes and their associated metrics were sourced from previously published large-scale exome sequencing data (Rolland et al., 2023). To visualize the intersection of genetic risk and fold change (FC) transcriptomic dysregulation, BETA values were plotted against log_2_FC values for both BMI and ASD datasets to highlight significant targets.

Transcriptomic downregulation can further be translated into a functional genetic risk framework by approximating an effective loss-of-function (LOF) allele dosage from differential expression FC. In a standard genetic model, risk is defined as the product of the effect size and the variant dosage Risk = β * Dosage (Collister et al., 2022). We modelled the RNA expression FC—derived from DESeq2 log_2_FC results comparing deletion (*DEL*) to control (*CON*) samples—as a proxy for LOF dosage. Under the assumption that normal expression (FC = 1.0) is equivalent to a wild-type state (Dosage = 0), a 50% reduction (FC = 0.5) mimics a heterozygous LOF state (Dosage = 1), and complete absence of expression (FC = 0) mimics a homozygous LOF knockout (Dosage = 2), the effective dosage for downregulated transcripts can be calculated as Dosage = 2 - (2 * FC).

### Single-Cell Regulatory Network Analysis

Gene regulatory networks (regulons) were inferred using *SCENIC* (Single-Cell regulatory Network Inference and Clustering) (ver. 1.3.1) (Aibar et al., 2017). Co-expression modules were identified using *GRNBoost2* function, and false positives were filtered based on cis-regulatory motif analyses (cisTarget). The regulons were identified in *CON* and *DEL* populations separately. Neurodevelopmentally regulated target genes identified by (Chu et al., 2021) were included for each regulon.

### Expression Variation Analysis (EVA)

To quantify the differential heterogeneity of gene regulation across conditions, we performed Expression Variation Analysis (EVA) (Davis-Marcisak et al., 2019). Unlike standard differential expression tests that compare mean expression levels, EVA utilizes a multivariate statistical framework to assess differences in the variance of transcriptional profiles. We applied EVA to assess heterogeneity in two types of gene sets. One was regulons derived from SCENIC, and another one was random gene sets, generated by the online tool MOLBIOTOOLS (https://molbiotools.com/) with default setting. Any gene that expressed less than 5% of all cells were excluded for further EVA. In details, we applied EVA to the single-cell regulon activity scores derived from SCENIC. For progenitor cells, we calculated the variation in regulon activity within each biological condition and performed a robust statistical test to identify regulons with significantly perturbed heterogeneity (dysregulation) between conditions. This allowed us to characterize condition-specific changes in regulatory network stability and diversity. For random gene sets, we generated 100 random gene sets, and each set contain 100 different genes, the variance of transcriptional profiles in each random gene set were established.

## RESULTS

### Genomic structure of hIPSC lines

The hIPSC lines used for this study were subjected to single nucleotide polymorphism (SNP) analysis (S1). This confirmed that the hIPSC cells were male (XY) and heterozygous for a deletion spanning the *16p11.2* locus from *SPN* to *CORO1A* in the *16p11.2^+/-^* lines (orange shading) and not in control *16p11.2^+/+^* lines as previously reported (Tai et al., 2016). The most frequent genomic feature identified by SNP analysis was loss of heterozygosity (green shading) present in the ancestral GM8 line and preserved control and deletion lines. Outside the *16p11.2* locus there were a small number of CNVs most of which had emerged after divergence from GM8. There were a few homozygous deletions (red shading) and homozygous duplications (purple shading) however most did not span genes so are not expected to affect gene dosage. In conclusion the only genomic change consistently distinguishing the *16p11.2^+/+^* and *16p11.2^+/-^*lines used in this study is the *16p11.2* microdeletion itself confirming these lines as essentially isogenic and suitable for investigating the molecular consequences of the *16p11.2* microdeletion without genetic variation outwith the *16p11.2* locus as a confounding factor.

### Characterisation of hIPSC derived cells

hIPSC cells were differentiated for 80 days following the protocol of (Y Liu et al., 2013) summarised in (S2, A) and checked for the telencephalic transcription factor FOXG1 and the post-mitotic cytoplasmic neuronal marker TUJ1 proteins. The vast majority of *CON* and *DEL* cell nuclei cell nuclei were FOXG1^+^ (S2, B,D) and many had acquired a TUJ1^+^ post-mitotic neuronal fate (S2, C,E) confirming telencephalic neuronal identity. Single cell RNA sequencing of three *CON* (C1, C2, and C3) and three *DEL* (D1, D2, D3) cultures yielded 33,954 cells across the six samples of which 14,916 comprised empty droplets, doublets, and cells with high levels of mitochondrial or ribosomal reads that were removed leaving 19,038 high quality cell transcriptomes for further analysis. We clustered cells using UMAP (Fig 1A) revealing two distinct populations both expressing the telencephalic early IN markers *FOXG1* and *NKX2.1* and identified the larger population (12,713 cells) as strongly expressing IN progenitor markers (*VIM*, *CLU*, *FABP7*) while the remainder (6,235 cells) expressed markers (*DLX2*, *INSM1*, *ZFHX3*) of more differentiated IN precursor cells more weakly expressed in progenitors (Fig 2B) (Shi et al., 2021; Yu et al., 2021). A recent study categorised single cell transcriptomes obtained from GW 9-12 human fetal ganglionic eminences as IN progenitors (denoted P1,P2,P3,P4,P5,P6) and more differentiated IN precursors (denoted LGE,MGE,CGE) (Yu et al., 2021). Comparison between our *in vitro* cell types and these *in vivo* cell types showed that the clusters we had identified as *in vitro* IN progenitors strongly correlated with the *in vivo* IN progenitors classed as P2 and P3 (stronger red shading in Fig 1C) consistent with high expression of *VIM*, *CLU*, and *FABP7* (Shi et al., 2021; Yu et al., 2021). The remainder of the *in vitro* cell population correlated with more differentiated LGE, CGE, and MGE precursor cell classes in the Yu study (Fig 1C). In conclusion, we have obtained single cell transcriptomes from isogenic hIPSCs with or without the *16p11.2* microdeletion differentiated into an *in vitro* correlate of IN progenitor cells found in the human fetal GE.

**Figure 1.**
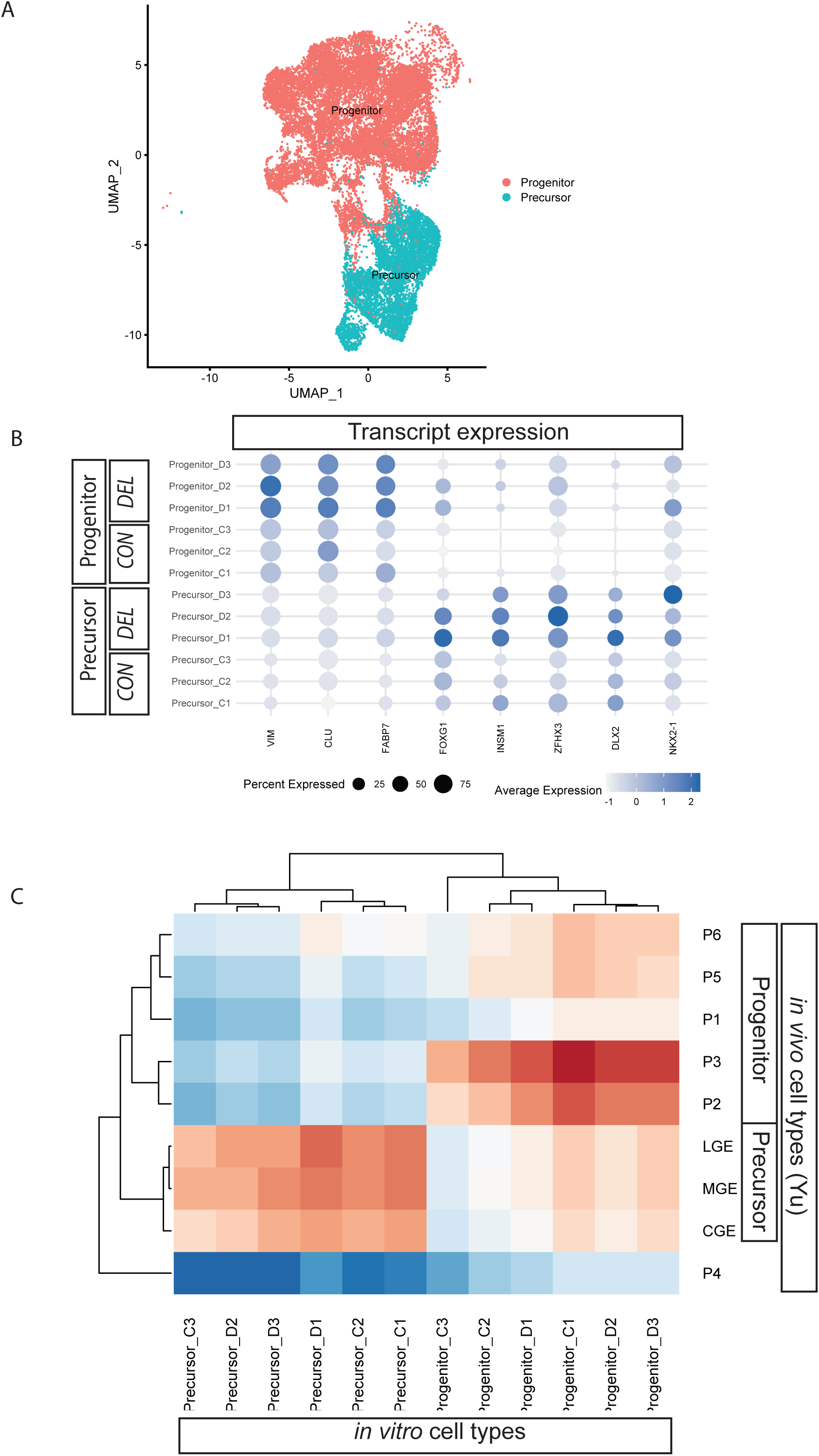
**(A)** UMAP showing dimensional reduction of the single cell transcriptomes for all samples of *CON* and *DEL* cells, each dot representing a single cell transcriptome, resolves into two main clusters identified as ‘progenitors’ (orange) and ‘precursors’ (turquoise). (B) Dot plot showing transcript expression in *CON* and *DEL* progenitor and precursor populations from all samples, for each transcript the dot indicates percent of expressing cells and average expression (see key) across cell populations. (C) Transcriptomic correlation between *in vitro* differentiated cell types (current study) and cell types from foetal human ganglionic eminences in vivo (Yu study) where P1-P6 are IN progenitors and LGE, MGE, and CGE are more differentiated IN precursor cells (Yu et al., 2021). Heatmap shading indicates strength of correlation between *in vitro* and *in vivo* cell types going from strongest (deep red), through white, to weakest (deep blue).

**FIGURE 2:**
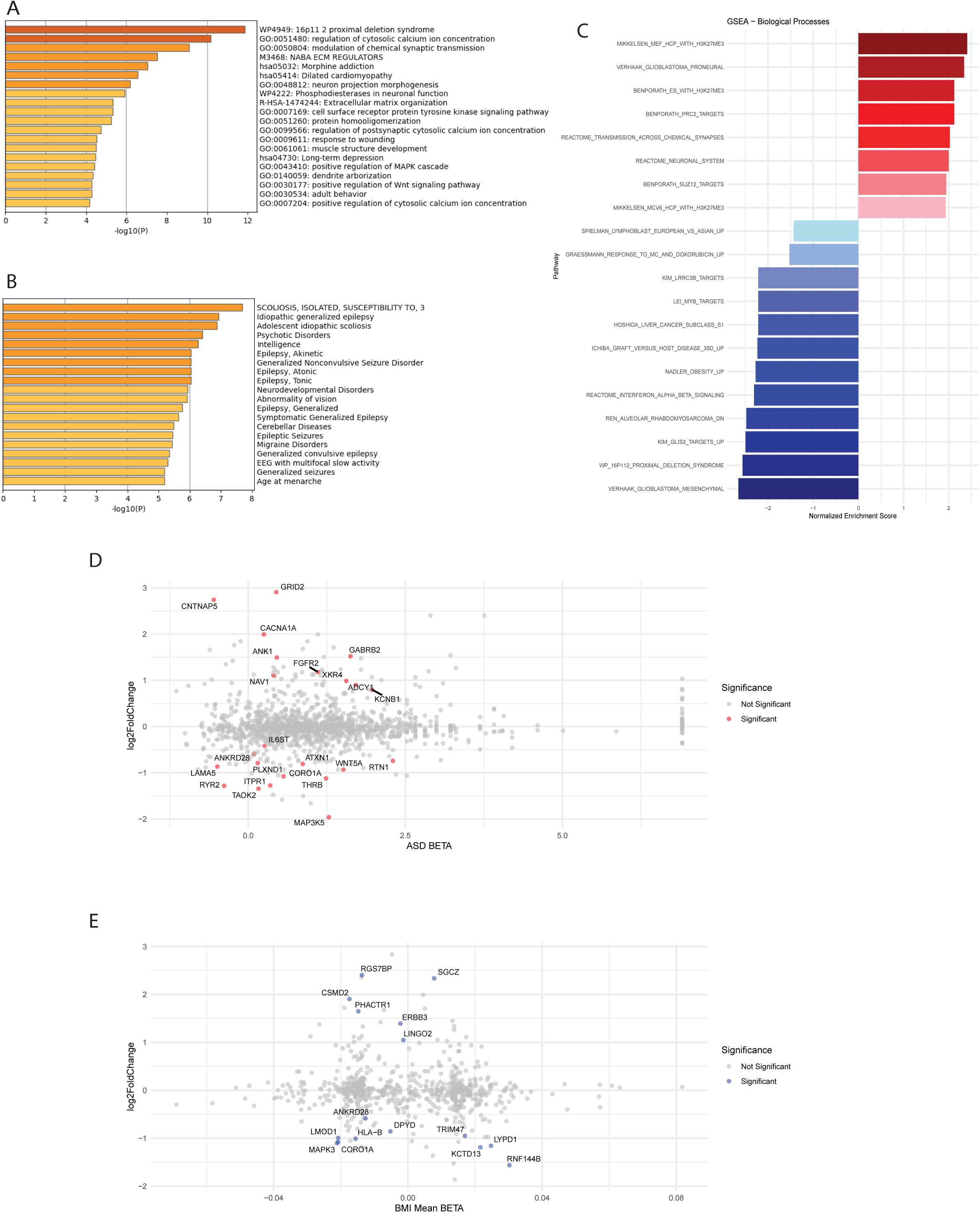
Analysis of differentially expressed genes between *CON* and *DEL* progenitor cells. (A) GO, (B) DisGeNET, (C) GSEA (D,E) Plots of log fold-change (Log_2_FC) transcript expression versus genetic risk (BETA value) associated with Autism (D) and BMI (E) for each gene where each corresponding transcript is a dot, only transcripts that are significantly differentially expressed (padj<0.05) between *CON* and *DEL* progenitors are coloured and their names indicated.

### Differential gene expression between *CON* and *DEL* progenitor populations

We next used DeSEq2 to identify differences in average ‘pseudo-bulk’ gene expression between *16p11.2^+/-^* and *16p11.2^+/+^* IN progenitors which we refer to as *DEL* (heterozygous for *16p11.2* microdeletion) and *CON* (control) respectively. After filtering out genes expressed in less than 5% of cells there were 185 significantly (DeSeq2 p_adj_ <0.05) differentially expressed genes (DEGs) in progenitor cells (S3). We used Metascape (Zhou et al., 2019) to perform gene ontology (GO) analysis showing that the most enriched term was *16p11.2* proximal deletion syndrome (Fig 2A) consistent with downregulation of *16p11.2* transcripts in *DEL* cells as has been observed in other studies. There was also some enrichment seen for other terms linked to calcium signalling, the synapse, signalling pathways, and brain development. DisGeNET showed enrichment of the DEGs in a number of human disease states with terms related to epilepsy and seizures, prominent features of *16p11.2* syndrome, being the most pronounced (Fig 2B).

As a complementary approach to examining differentially expressed genes we used gene set enrichment analysis (GSEA) (Subramanian et al., 2005) which ranks transcripts by their fold-change between cell populations and identifies gene-sets which are either enriched in up-regulated transcripts (positive normalised enrichment score) or in down-regulated transcripts (negative normalised enrichment score) regardless of whether they are called as significantly dysregulated by DeSEq2. In progenitor cells the majority of top terms with highest positive enrichment were gene-sets linked to chromatin biology (MIKKELSEN_MEF_HCP_WITH_H3K27ME3, BENPORATH_ES_WITH_H3K27ME3, BENPORATH_PRC2_TARGETS, and BENPORATH_SUZ12_TARGETS, MIKKELSEN_MCV6_HCP_WITH_H3K27ME3), cancer related terms were present in both positively (VERHAAK_GLIOBLASTOMA_PRONEURAL) and negatively (HOSHIDA_LIVER_CANCER_SUBCLASS_S1 and VERHAAK_GLIOBLASTOMA_MESENCHYMAL) enriched. The *16p11.2* locus transcripts (WP_16P112_PROXIMAL_DELETION_SYNDROME) were negatively enriched and, interestingly, so was an obesity related term (NADLER_OBESITY_UP) (Fig 2C).

The *16p11.2* microdeletion increases the odds of developing autism and obesity (body mass index (BMI) > 30 kg/m^2^) the most in *16p11.2* microdeletion carriers, however, unlike epilepsy, neither features prominently in our DisGeNET results (Fig 2B). We took advantage of publicly available data from from genome wide association studies (GWAS) reporting the association between single nucleotide polymorphism (SNP) loci and either autism as a binary trait (Rolland et al., 2023) or body mass index (BMI) as a continuous trait defined as weight in kilograms divided by height in meters squared from UK BIOBANK (Bycroft et al., 2018). While a SNP associated with altered odds of developing a trait is itself unlikely to be directly causing the condition it may be genetically linked by being in close proximity to the causal allele in the genomic DNA making mutations in genes near the SNP strong candidates. For GWAS studies the ‘effect size’ or β (BETA), calculated from the odds ratio (OR) with BETA = log_e_OR indicates the contribution of each allele to the condition with increasingly positive BETA indicating increased risk, BETA = 0 indicating no risk, while negative BETA values indicate a risk reducing (protective) effect (Collister et al., 2022). To visualise the relationship for each gene between transcript dysregulation in progenitor cells and GWAS association with either autism (S4) or obesity (S5) we plotted the Log2FC value against the BETA value for autism (Fig 2D) or BMI (Fig2E). While the consequences of up-regulation are more difficult to predict down-regulated transcripts in *DEL* progenitors may approximate loss of function that scales with increasing down-regulation (see methods) and the combined effect of transcripts down-regulated about 2-fold (Log_2_FC ≈ -1) could have polygenic effects analogous to loss of function heterozygosity for multiple risk loci.

### Gene set transcript level variability between *CON* and *DEL* progenitor populations

Phenotypic variation in *16p11.2* heterozygotes prompted us to next investigate gene expression variability between cells harbouring the *16p11.2* microdeletion. We turned to Expression Variation Analysis (EVA) (Davis-Marcisak et al., 2019) which has been used to identify increased transcript level heterogeneity in tumour cells compared to their non-tumorous counterparts. This method calculates an EVA statistic for a gene set in a group of cells, with higher EVA values indicating increased variability. We calculated the EVA score for each gene-set for the progenitor cells in each sample *CON* (C1,C2,C3) and *DEL* (D1,D2,D3) separately, the average EVA value for the CON and DEL samples, and the difference between average CON and DEL EVA averages (EVA(*DEL-CON*)) to provide a measure of differences in gene expression variability between *DEL* and *CON* samples with positive EVA(*DEL-CON*) indicating greater variability in *DEL* samples for that gene set. To provide a measure of the degree of differential gene expression in each gene-set we divided the number of transcripts more than 2-fold up- or down- regulated (1<Log2FC<-1) (D) by the number of transcripts in the gene-set (N) to give the D/N value for each regulon.

### Regulon gene-sets exhibiting greater variability in *DEL* progenitors show increased enrichment of genes linked to the cell cycle and cancer

Regulons describe genes functionally connected by transcriptional regulation and each comprise a transcription factor (the HubTF) and the genes it transcriptionally regulates (the targets). We used SCENIC (Aibar et al., 2017) to identify 94 regulons in *DEL* (S6) and *CON* (S7) samples and calculated their EVA scores (S8, S9) in *DEL* and *CON* progenitors. This revealed a significant difference (n=3, paired t-test p=3.37x10^-7^) indicating that *16p11.2* genotype affects gene expression variability. The distribution of EVA(*DEL-CON*) values shows a clear bias towards positive values (Fig 3A) indicating more gene expression variability in the *DEL* progenitor population. There is a weak positive correlation (correlation coefficient = +0.25) between D/N and EVA(*DEL-CON*). We ranked the 94 regulons by EVA(*DEL-CON*) value and divided them equally into quintiles Q1 – Q5 (indicated on Fig 3B). We then input all the target genes in each quintile to Metascape (Zhou et al., 2019) to relate the degree of gene expression variability to enrichment of gene sets associated with biological processes (gene ontology (GO) analysis) and disease associated genes (DisGeNET) to these quintiles.

**FIGURE 3:**
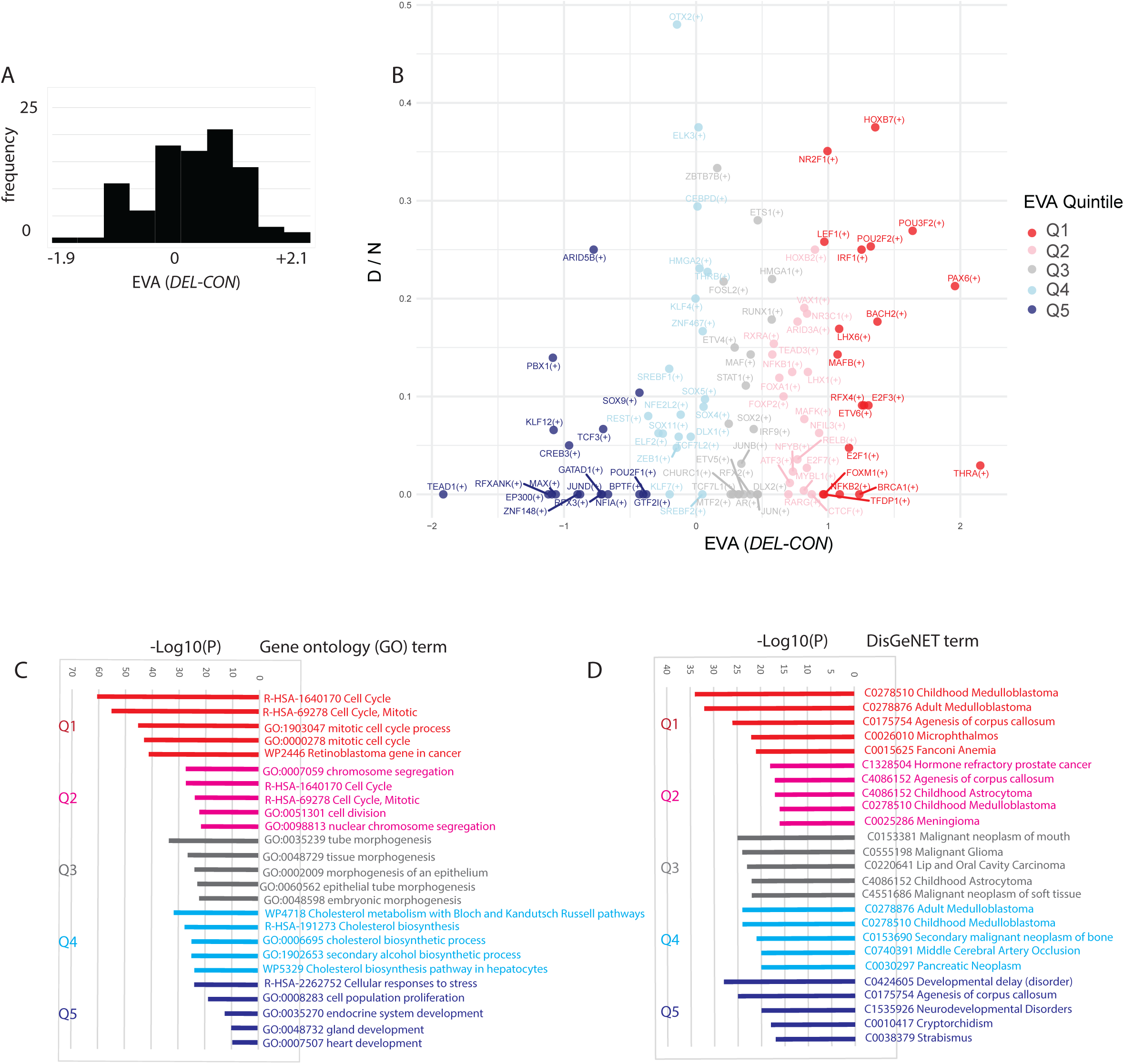
(A,B) Comparative expression variation analysis (EVA) of 94 regulon gene sets in *CON* and *DEL* progenitor cell populations. EVA(*DEL-CON)* is the difference between average EVA scores of all *DEL* and *CON* progenitor samples with a positive score indicating increased gene expression variability in *DEL* samples. (A) The distribution of EVA(*DEL-CON*) values shows a bias to positive scores. (B) Relationship between the D/N value, the proportion of target transcripts in each regulon up – or down – regulated more than 2 fold (1<Log2FC<-1), and the EVA(*DEL-CON*) value for each regulon. The hubTF for each regulon indicated (eg THRA1(+) is the regulon with THRA as the HubTF). Q1-Q5 indicate the quintiles used for gene ontology and DisGeNET with Q1 representing the regulons with the greatest increased variability in *DEL* samples. (C) Gene ontology (GO) analysis of the ‘target’ genes of the regulons in each of the quintiles. (D) DisGeNET analysis of the ‘target’ genes of the regulons in each of the quintiles. In C,D the top 5 enriched terms for each quintile and their -log_10_P values indicated.

GO terms related to cell division (‘cell cycle’, ‘DNA replication’, ‘regulation of cell cycle phase’, ‘S-phase’, cell population proiferation’, mitotic cell cycle process’, ‘chromosome segregation’) were represented across all quintiles with enrichment particularly pronounced (highest - log10(P) value indicating higher proportion of genes from a GO term present in the quintile gene-set) in Q1, the quintile containing regulons with the greatest increased variability in *DEL* progenitors, with a trend towards lower significance in quintiles containing successively less variable regulons. This suggests that regulons exhibiting the greatest gene expression variation in *DEL* progenitor cells compared to *CON* cells are functionally involved in cell division.

DisGeNET terms associated with cancers (‘Childhood meduloblastoma’, ‘adult medulloblastoma’, ‘squamous cell carcinoma of lung’) were prominently enriched in genes from Q1, the regulons exhibiting most increased variation in *DEL* compared to *CON* progenitors, and perhaps this is not surprising given the well known role of cell cycle dysregulation in cancers. ‘Agenesis of the corpus callosum’, a sensitive neurodevelopmental indicator, is similarly represented across quintiles while DisGeNET terms associated with neurodevelopmental disorders (‘Developmental delay (disorder)’, ‘Neurodevelopmental disorders’) were most significantly enriched in Q5 representing regulons with decreased variation in *DEL* compared to *CON* progenitors. Together this suggests that abnormal gene expression variation may each correlate with phenotypic outcomes.

### Increased gene-set expression variability in DEL progenitors extends to randomly generated gene-sets

To investigate whether the increased regulon gene expression variation in *DEL* progenitors applied more generally we generated 100 random gene sets each comprising 100 genes (S10) and subjected them to the same EVA analysis described above for the regulon gene sets (S11). There was a significant difference in EVA values between *CON* and *DEL* progenitors (n=3, paired t-test P=5.94x10^-25^) with EVA(*DEL-CON*) values showing a strong bias towards positive values (Fig 4A) indicating increased gene expression variability in DEL progenitor populations even in unrelated sets of genes. As previously observed for the regulons there was a weak negative correlation (correlation coefficient = -0.13) between EVA(*DEL-CON*) and D/N values (Fig 4B). We next ranked the 100 random gene sets according to EVA(*DEL-CON*) values and divided them into quintiles of 20 gene-sets each to look for enrichment of GO and DisGeNET terms. In contrast to the regulons, and not particularly surprisingly given the gene-sets were randomly generated with no functional connection, enrichment was much more modest and less obviously linked to relevant processes (Fig 4C) or diseases (Fig 4D) than observed for the regulons (Fig 3C,D) also providing a control for the substantial enrichment we observed in the most variable regulons.

**FIGURE 4:**
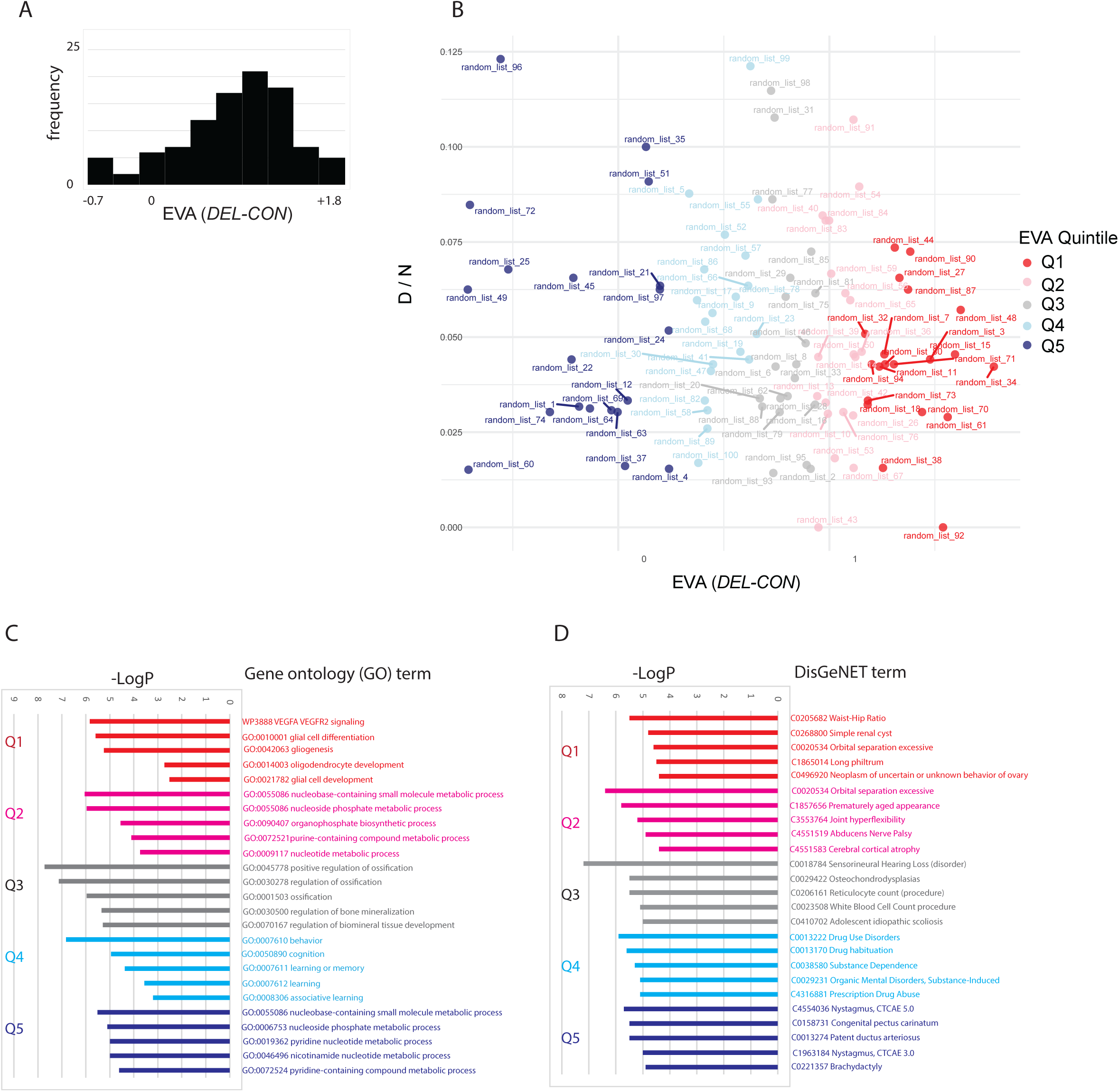
(A,B) Comparative expression variation analysis (EVA) of 100 random gene-sets of 100 genes in *CON* and *DEL* progenitor cell populations. EVA(*DEL-CON)* is the difference between average EVA scores of *DEL* and *CON* progenitor samples with a positive score indicates increased gene expression variability in *DEL* samples. (A) The distribution of EVA(*DEL-CON*) values shows a bias to positive scores. (B) Relationship between differential gene expression within the randon gene-sets (D/N) value, the proportion of transcripts in each gene-set up – or down – regulated more than 2 fold (1<Log2FC<-1), and the EVA(*DEL-CON*) value for each gene-set. Q1-Q5 indicate the quintiles used for gene ontology and DisGeNET with Q1 representing the gene-sets with the greatest increased variability in *DEL* samples. (C) Gene ontology (GO) analysis of the gene-sets in each of the quintiles. (D) DisGeNET analysis of the gene-sets in each of the quintiles. In C,D the top 5 enriched terms for each quintile and their -log_10_P values indicated.

## Discussion

The *16p11.2* microdeletion is associated with the symptoms of *16p11.2* syndrome with considerable variation between individual carriers. In the current study we investigate the transcriptional consequences of the *16p11.2* microdeletion for *in vitro* correlates of ventral telencephalic IN progenitor cells in the first trimester at gestational weeks 10 to 12.

Examination of differentially expressed genes (DEGs) revealed that even at the IN progenitor stage enrichment of disease genes (DisGeNET) associated with epilepsy, a condition found in 20% of *16p11.2* microdeletion carriers, dominated while prominent among gene ontology (GO) terms enriched in the DEGs were calcium signalling, the synpase, cell signalling, and the extracellular matrix. This resembles other studies where IPSC cells with and without the *16p11.2* microdeletion are differentiated into other types of neural progenitors and their transcriptomes compared revealing hundreds of DEGs, each of which is relatively modestly up or down regulated, with a common theme of involvement in cell signalling and neurodevelopmental disorders(Blumenthal et al., 2014; Connacher et al., 2022; Roth et al., 2020; Urresti et al., 2021). This raises the question of how dysregulation in progenitor cells of genes generally associated with more differentiated cell structures, such as the synapse, contribute to the *16p11.2* phenotype. In contrast, gene set enrichment analysis (GSEA), a method based solely on the degree of up or down regulation and not statistical significance, found enriched terms linked to cancer and histone modification. We also noticed that there were quite large numbers of modestly up- or down-regulated transcripts of genes associated with autism or obesity in GWAS studies. GWAS identifies association between genomic DNA and disease states and is agnostic about the site or time of action. Finding that genes associated with autism or obesity are dysregulated so early in the IN developmental trajectory raises the possibility of a cumulative polygenic effect on progenitor phenotype caused by their dysregulation that has collateral effects reaching into post-natal life. Alternatively similar transcriptomic variability may well also manifest in more differentiated cells with more direct effects. Clearly this needs to be investigated in much more depth.

In a recent study we differentiated isogenic human *CON* (*16p11.2^+/+^*) and *DEL* (*16p11.2^+/-^*) IPSC cells into ventral telencephalic organoids and found that the *DEL* organoids exhibited a phenotype of increased variability in organoid size, cell cycle kinetics, and accelerated maturation (Fetit et al., 2023). This suggested that increased phenotypic variability in organoid culture is a consequence of reduced dosage of the *16p11.2* locus itself. This all prompted us to ask whether focusing on gene expression variability rather than solely on differential expression would give us additional insight into the *16p11.2* phenotype and used single cell expression variation analysis (EVA) to compare variation in the expression of gene-sets between individual cells in the *CON* and *DEL* progenitor cell populations (Davis-Marcisak et al., 2019). We examined 94 regulons identified by SCENIC in our single cell transcriptomes and found a strong bias towards increased gene expression variation in samples harbouring the *16p11.2* microdeletion. The most variable 19 regulons (Quintile 1) presented the strongest enrichment in GO terms linked to the cell cycle and DisGeNET terms associated with cancer. The significant overlap between genes associated with cancer and those associated with autism and other neurodevelopmental conditions has been interpreted as indicating common underlying molecular mechanisms (Forés-Martos et al., 2019). Directly relevant to the current study the *16p11.2* microdeletion has been shown to increase the odds of developing neuroblastoma 13 fold, comparable to increased odds for other *16p11.2* symptoms, indicating a dual function in cancer and neurodevelopment (Egolf et al., 2019). While we previously found that ventral telencephalic organoids harbouring the *16p11.2* microdeletion generally presented more variable phenotypes than their wild-type counterparts this study did not address cell-cell variation (Fetit et al., 2023). Our finding here that the *16p11.2* microdeletion increases gene expression heterogeneity between cells is reminiscent of cancers where EVA analysis of single cell transcriptomes reveals increased gene expression variation in large numbers of tumour samples compared to their non-tumerous counterparts (Davis-Marcisak et al., 2019). Increased cell-cell variability in regulon transcript levels associated with the cell-cycle extends our earlier finding of increased organoid-organoid cell-cycle variability (Fetit et al., 2023). Less expected was that this effect was much more widely distributed across the genome as randomly selected gene sets also showed a strong bias to increased gene expression variation between cells harbouring the *16p11.2* microdeletion. Together this suggests the possibility that tumours and neurodevelopmental conditions not only share overlapping sets of genes but also gene expression heterogeneity as a possibly underpinning phenotype.

Our finding that dosage of the *16p11.2* locus has a widespread influence on cell-cell transcriptional variation for thousands of genes scattered about the human genome poses the question of the impact. Given the highly constrained boundaries for cell function it is easy to imagine that global transcript instability, with knock-on consequences for bio-molecule interactions, has far reaching consequences. In addition to cancer biology where molecular heterogeneity has long been associated with disease progression it has more recently been reported that gene expression variation associates with neurodevelopmental conditions. A recent study finds increased gene expression variability in human IPSC derived neurons harbouring trisomy 21 or *CHD8* mutations, both risk factors for neurodevelopmental conditions(Upadhya et al., 2025). Trisomy 21 is a polygenic risk factor increasing dosage of hundreds of genes on chromosome 21 so it is unknown which underlie the gene expression variation phenotype. The *CHD8* mutation is a monogenic risk factor and *CHD8* mutations likely impact on gene expression variability stemming from its role in chromatin remodelling (Wang et al., 2017). GSEA analysis (current study) identified enrichment and depletion of DEGs associated with chromatin in *DEL* progenitors consistent with the idea that chromatin disruption is a factor in the *16p11.2* phenotype, furthermore the *16p11.2* locus includes *HIRIP3* and *PAGR1* which have functions in chromatin biology so are well placed, like *CHD8*, to exert widespread effects on the stability of gene expression when their dosage is reduced by the *16p11.2* microdeletion.

One possible consequence of increased transcript expression variability of a regulon is disruption of the processes normally controlled by that regulon. Indeed, several of the most disorganised regulons are controlled by TFs (the hub TFs), that are themselves associated with aspects of *16p11.2* syndrome. Thyroid hormone (TH) acts by binding to its receptor encoded by *THRA* enabling it to activate gene expression by binding to thyroid response elements (TRE) in target gene promotors and dysregulation of thryroid signalling is associated with a wide variety of symptoms with *THRA* mutations identified as risk factors for obesity and autism (Fernández-Real et al., 2013; Kalikiri et al., 2017; Yuen et al., 2015). Obesity has also been associated with altered chromatin accessibility of TRE sites to THRA protein in adipocytes suggesting that *THRA* phenotypes can be mediated by dysregulating THRA activated transcription without directly affecting THRA itself (Zhu et al., 2025). Cilia are cellular antenna essential for many signalling pathways and mutations affecting cilia components predispose to symptoms overlapping with *16p11.2* syndrome, for example obesity and autism, with disruption to cilia reported in *16p11.2* microdeleted cells (Harris et al., 2021; Migliavacca et al., 2015; Schwartz et al., 2011). *RFX4*, along with several other *RFX* transcription factors, regulates transcription of cilia genes and RFX4 binds to promoters and regulates *POU3F2* and *NEUROD2* proneural gene transcription in human neural progenitors (Choi et al., 2024). Notably POU3F2 is the HubTF for a regulon with both one of the highest proportions of differentially expressed target transcripts and one of the highest relative transcript level variability in our *16p11.2* heterozygous progenitors and loss of function *POU3F2* alleles associate with obesity and neurodevelopmental disorders (Schönauer et al., 2023). While *PAX6* mutations were first known for causing the ocular condition aniridia and there is accumulating evidence that heterozygosity for *PAX6* loss of function also affects brain anatomy and function (Abouzeid et al., 2009; Bamiou et al., n.d.; Grant et al., 2021; Hanish et al., 2016; Maekawa et al., 2009; Manuel et al., 2015; Yogarajah et al., 2016). Elevated *E2F1* levels have been associated with obesity in humans and there is evidence for *E2F1* acting both in adipose cells and in the hypothalamic appetite regulating cells to regulate metabolism and appetite (Denechaud et al., 2017; Haim et al., 2015; Lu et al., 2013). *NR2F1* is expressed in IN progenitors and associated with autism (Zhang et al., 2020). Overall the approach of using variation as a criteria over and above sole focus on differential gene expression has yielded regulons with known clinical impact that merit further investigation. Given the widespread expression of the *16p11.2* locus it will be interesting to see whether this enhanced molecular variability phenotype of the *16p11.2* locus applies more broadly and investigate the mechanism and phenotypic consequences in more depth.

## Data Availability

All data produced in the present study are available upon reasonable request to the authors

## Supplementary figures/ tables

**S1 SNP microarray:** Single nucleotide polymorphism (SNP) analysis of IPSC lines showing genomic copy number variation. GM8 is the ancestral line used to derive the control [FACS52 (C1), FACS53 (C2 and C3)] and *16p11.2* microdeletion [DELD5 (D1), DELH7 (D3), and DELC5 (D2)] samples as described previously (Tai et al., 2016). The key indicates different types of copy number variation and the position of the 16p11.2 locus on chromosome 16 is indicated confirming a heterozygous deletion in DEL and not CON lines. No other copy number variation consistently partitions between *CON* and *DEL* lines confirming they are overwhelmingly isogenic except for the *16p11.2* locus itself.

**S2 IPSC culture:** (A) Summary of culture protocol used to differentiate *CON* and *DEL* isogenic IPSC cells (Y Liu et al., 2013)for 80 days prior to dissociation for single cell RNA sequencing. (B-D) Immunofluorescence for FOXG1 and TUJ1 on a CON (B,C) and DEL (D,E) culture after 80 days in cuture. B,D shows the FOXG1 signal only while C,E show the TUJ1, FOXG1 and DAPI channels merged.

**S3 DESeq2 DEGs:** Differential gene expression between *CON* and *DEL* progenitors from DESeq2 analysis. First column indicates transcript. For each transcript (row): pct_exp_CON_Prog and pct_exp_DEL_Prog is the percentage of cells transcript expression is detected in CON and DEL respectively; baseMean_Prog is overall transcript expression level; Log2FoldChange is log (base 2) of the differential transcript expression fold change in *CON* relative to *DEL* progenitors with negative values indicating reduced expression in DEL compared to CON. LfcSE is the standard error for the Log2FoldChange and pvalue and padj are the unadjusted and adjusted p values respectively.

**S4 autism BETA:** First column indicates transcript. For each transcript (row) the odds ratios (autism_OR) and the beta (BETA) value using log (base e) OR giving the effect size for association with autism (see Methods).

**S5 BMI BETA:** First column indicates transcript. For each transcript (row) the BMI_mean_BETA is the effect size for association with BMI calculated by averaging the individual BETA values of SNPs in a 32.5 kilobase pair window encompassing each gene (see Methods).

**S6 SCENIC regulons DEL:** First column indicates the HubTF for each regulon and the second column indicates it’s transcriptional targets as identified by SCENIC in the *DEL* single cell transcriptomes (see Methods).

**S7 SCENIC regulons CON:** First column indicates the HubTF for each regulon and the second column indicates it’s transcriptional targets as identified by SCENIC in the *CON* single cell transcriptomes (see Methods).

**S8 EVA for regulons *DEL*:** Regulons identified in the *DEL* samples with their EVA scores in all the samples. First column indicates the hubTF for each regulon. For each row N_Prog is the number of expressed transcripts in each regulon; D_Prog is the number of transcripts expressed more than 2-fold differently between CON and DEL samples; D-N_Prog is D divided by N (the proportion of transcripts in each regulon expressed more than 2-fold differently between *CON* and *DEL* samples); and EVA values for each *CON* (Prog_C1, Prog_C2, and Prog_C3) and *DEL* (Prog_D1, Prog_D2, and Prog_D3) progenitor sample for each regulon (see Methods).

**S9 EVA for regulons *CON*:** Regulons identified in the *CON* samples with their EVA scores in all the samples. First column indicates the hubTF for each regulon. For each row N_Prog is the number of expressed transcripts in each regulon; D_Prog is the number of transcripts expressed more than 2-fold differently between CON and DEL samples; D-N_Prog is D divided by N (the proportion of transcripts in each regulon expressed more than 2-fold differently between CON and DEL samples); and EVA values for each *CON* (Prog_C1, Prog_C2, and Prog_C3) and *DEL* (Prog_D1, Prog_D2, and Prog_D3) progenitor sample for each regulon (see Methods).

**S10 Random gene sets:** Each row shows a list of 100 randomly selected genes. The first column shows the identifier used to refer to each gene set.

**S11 EVA random gene sets:** EVA scores for random gene-sets in all the samples. First column indicates the 100 random gene set identifier. For each row D_N_Prog is the proportion of transcripts expressed more than 2-fold differently between *CON* and *DEL* samples expressed transcripts in each set of 100 random genes; and EVA values for each *CON* (Prog_C1, Prog_C2, and Prog_C3) and *DEL* (Prog_D1, Prog_D2, and Prog_D3) progenitor sample for each random gene set (see Methods).

## Contributions and acknowledgements

IQ performed cell culture, YY performed bioinformatic analysis, TP designed the study and wrote the paper. We thank James Gusella and Derek Tai at the Molecular Neurogenetics Unit, Harvard Medical School (Boston, MA, USA), for providing the cell lines used in this study. RNA sequencing was performed by Edinburgh Genomics. This work was funded by the Simons Initiative for the Developing Brain (SFARI - 529085) and Biotechnology and Biological Sciences Research Council (BB/M00693X/1).

## Notes

### Competing Interest Statement

The authors have declared no competing interest.

### Author Declarations

The study used ONLY openly available human data that were originally located at UK Biobank OR published in the scientific literature cited in the manuscript.

