## Supplementary figures and images for "The *16p11.2* microdeletion enhances gene expression variability between human IPSC derived forebrain interneuron progenitor cells in culture"

### S1

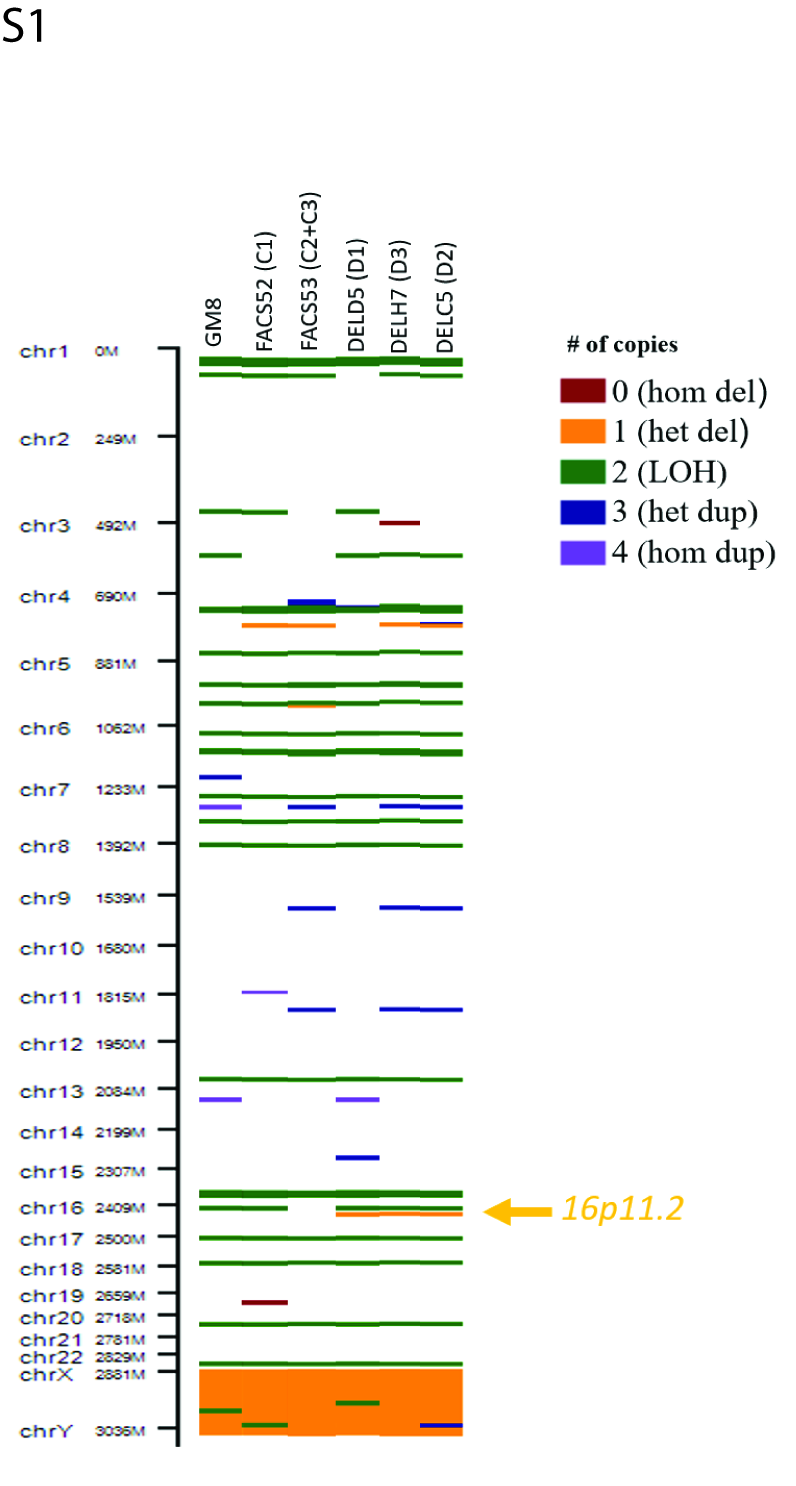

### S2

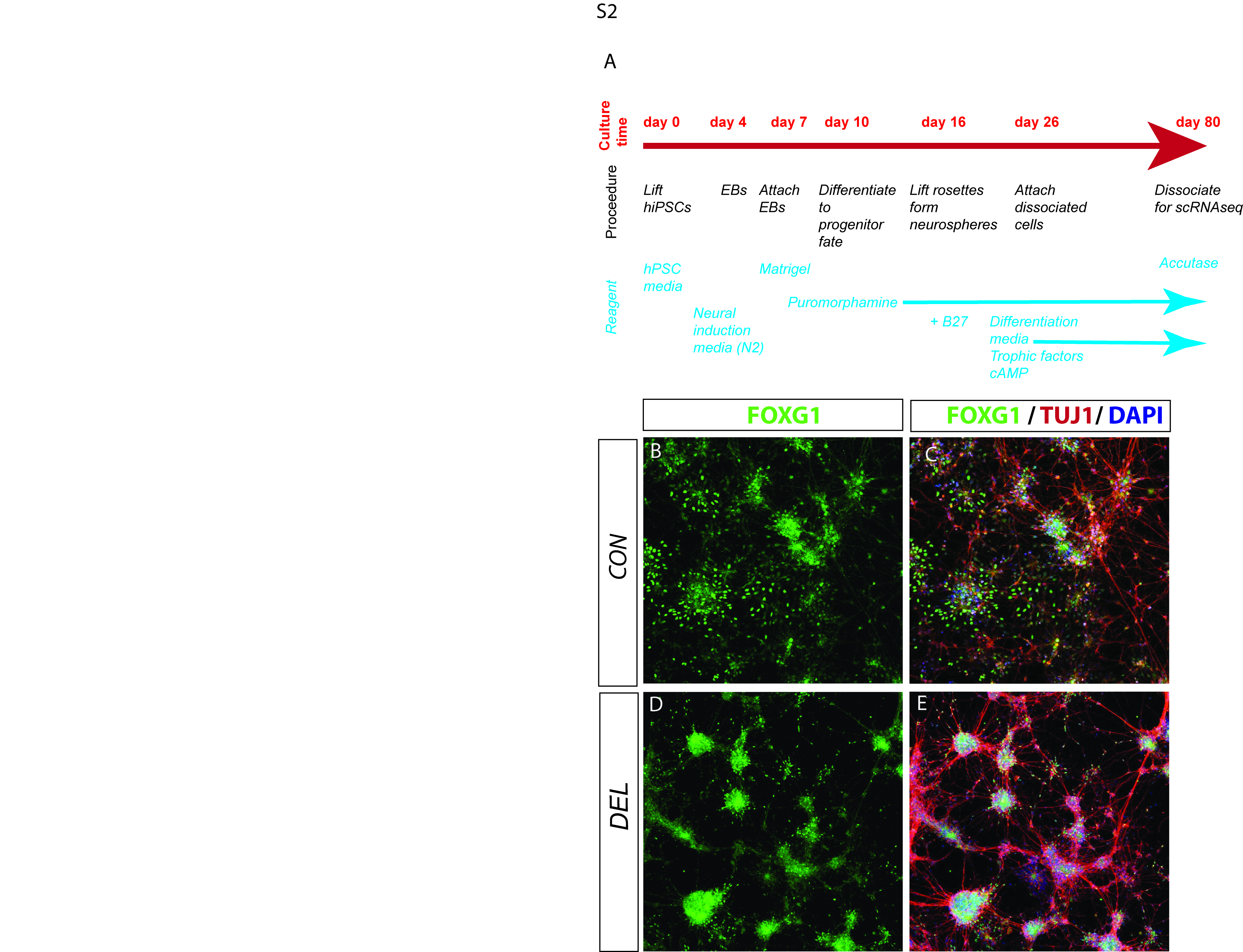
